# Safety and immunogenicity of inactivated whole virion vaccine CoviVac against COVID-19: a multicenter, randomized, double-blind, placebo-controlled phase I/II clinical trial

**DOI:** 10.1101/2022.02.08.22270658

**Authors:** Aydar A Ishmukhametov, Aleksandra A Siniugina, Nadezhda V Yagovkina, Vladimir I Kuzubov, Konstantin A Zakharov, Viktor P Volok, Maria S Dodina, Larissa V Gmyl, Natalya A Korotina, Rostislav D Theodorovich, Yulia I Ulitina, Andrey A Tsaan, Tatiana V Pomaskina, Anna V Kalenskaya, Irina V Solovjeva, Elena V Tivanova, Larissa Y Kondrasheva, Antonina A Ploskireva, Vasiliy G Akimkin, Ksenia A Subbotina, Georgy M Ignatyev, Anastasia K Korduban, Elena Y Shustova, Ekaterina O Bayurova, Alla S Kondrashova, Darya V Avdoshina, Anastasia N Piniaeva, Anastasia A Kovpak, Liliya P Antonova, Yulia V Rogova, Anna A Shishova, Yury Y Ivin, Svetlana E Sotskova, Konstantin A Chernov, Elena G Ipatova, Ekaterina A Korduban, Liubov I Kozlovskaya, Ilya V Gordeychuk

**Author notes:** **Corresponding author:** Ilya Gordeychuk; address: premises 8, bldg. 1, Village of Institute of Poliomyelitis, Settlement “Moskovskiy”, Moscow 108819, Russia; add. 1035. **Author contributions:** Study concept and design — Aydar A Ishmukhametov, Aleksandra A Siniugina, Andrey A Tsaan, Ekaterina A Korduban, Georgy M Ignatyev, Liubov I Kozlovskaya Ilya V Gordeychuk; acquisition of data — Nadezhda V Yagovkina, Vladimir I Kuzubov, Konstantin A Zakharov, Viktor P Volok, Maria S Dodina, Larissa V Gmyl, Natalya A Korotina, Rostislav D Teodorovich, Yulia I Ulitina, Andrey A Tsaan, Tatiana V Pomaskina, Anna V Kalenskaya, Irina V Solovjeva, Elena V Tivanova, Larissa Y Kondrasheva, Antonina A Ploskireva, Vasiliy G Akimkin, Ksenia A Subbotina, Georgy M Ignatyev, Anastasia K Korduban, Elena Y Shustova, Ekaterina O Bayurova, Alla S Kondrashova, Darya V Avdoshina, Anastasia N Piniaeva, Anastasia A Kovpak, Lilia P Antonova, Yulia V Rogova, Anna A Shishova, Yury Y Ivin; analysis and interpretation of data — Aydar A Ishmukhametov, Ekaterina Bayurova, Liubov I Kozlovskaya, Ilya V Gordeychuk; drafting of the manuscript — Liubov I Kozlovskaya, Ilya V Gordeychuk, Alla S Kondrashova, Darya V Avdoshina; critical revision of the manuscript for important intellectual content — Aydar A Ishmukhametov, Liubov I Kozlovskaya, Ilya V Gordeychuk; statistical analysis — Andrey A Tsaan, Ksenia A Subbotina, Yulia I Ulitina, Liubov I Kozlovskaya; obtained funding — Aydar A Ishmukhametov; administrative, technical, or material support — Aleksandra A Siniugina, Anastasia K Korduban, Anastasia N Piniaeva, Ekaterina A Korduban, Elena G Ipatova, Georgy M Ignatyev, Konstantin A Chernov, Konstantin A Zakharov, Nadezhda V Yagovkina, Svetlana E Sotskova, Tatiana V Pomaskina, Vladimir I Kuzubov, Antonina A Ploskireva, Vasiliy G Akimkin; study supervision — Aydar A Ishmukhametov, Aleksandra A Siniugina, Andrey A Tsaan, Ekaterina A Korduban. (A.A.I.), (A.A.S.), (V.P.V.), (M.S.D.), (L.V.G.), (N.A.K.), (R.D.T.), (G.M.I.), (A.K.K.), (E.Y.S.); (E.O.B.), (A.S.K.), (D.V.A.), (A.N.P.), (A.A.K.), (L.P.A.), (Y.V.R.), (A.A.S.), (Y.Y.I.); (S.E.S.), (K.A.C), (E.G.I.), (E.A.K.), (L.I.K.), (I.V.G.). (N.V. Y.). (V.I.K.). (K.A. Z.). (A.A.T.); (Y.I.U). (T.V.P.). (A.V.K.), (I.V.S.), (E.V.T.), (L.Y.K.), (A.A.P.), (V.G.A.). (K.A.S.).

## Abstract

We present the results of a randomized, double-blind, placebo-controlled, multi-center clinical trial of the tolerability, safety, and immunogenicity of the inactivated whole virion concentrated purified coronavirus vaccine CoviVac in adult volunteers aged 18-60.

Safety of the vaccine was assessed in 398 volunteers who received two doses of the vaccine (n=298) or placebo (n=100). The studied vaccine has shown good tolerability and safety. No deaths, serious adverse events (AE), or other significant AE related to vaccination have been detected. The most common AE in vaccinated participants was pain at the injection site (p<0.05).

Immunogenicity assessment was performed in 167 volunteers (122 vaccinated and 45 in Placebo Group) separately for the participants who were anti-SARS-CoV-2 nAB negative (69/122 in Vaccine Group and 28/45 in Placebo Group) or positive (53/122 in Vaccine Group and 17/45 in Placebo Group) at screening.

At Day 42 after the first immunization the seroconversion rate in participants who were seronegative at screening was 86.9% with average the geometric mean neutralizing antibody (nAB) titer of 1:20. Statistically significant (p<0.05) increase of IFN-γ production by peptide-stimulated T-cells was observed at Days 14 and 21 after the first immunization.

In participants who were seropositive at screening but had nAB titers below 1:256 the rate of 4-fold increase in nAB levels was 85.2%, while in the participants with nAB titers >1:256 the rate of 4-fold increase in nAB levels was below 45%. For the participants who were seropositive at screening the second immunization did not lead to a significant increase in nAB titers.

In conclusion, inactivated vaccine CoviVac has shown good tolerability and safety, with 86.9% seroconversion rates in participants, who were seronegative at screening. In participants who were seropositive at screening and had nAB titers below 1:256, a single immunization lead to a 4-fold increase in nAB levels in 85.2% cases.

## INTRODUCTION

The ongoing pandemic of coronavirus disease 2019 (COVID-19), which is caused by severe acute respiratory syndrome coronavirus 2 (SARS-CoV-2), has claimed more than five million lives as of November 2021 (WHO, 2021). In order to ensure the availability of the vaccines against COVID-19 to every country in the world, all existing vaccine platforms and manufacturing capacities were employed, including genetic (Baden et al., 2021; Polack et al., 2020), vectored (Falsey et al., 2021; Logunov et al., 2021), subunit (Heath et al., 2021) and inactivated (Al Kaabi et al., 2021; Ella et al., 2021; Zakarya et al., 2021; Zhang et al., 2021) vaccines etc.

Despite the strong immunogenicity and high short-term efficacy of the abovementioned approaches, it has been shown that post-vaccination antibody levels decrease over time (Crawford et al., 2021; Levin et al., 2021), which has led to implementation of annual and even semiannual booster vaccinations in unfavorable epidemiological conditions in several countries, including Russia, setting even greater demands for the safety profile of the vaccines used.

Despite generally lower post-vaccination neutralizing antibody (nAB) levels as compared to vectored and mRNA vaccines, inactivated vaccines have been instrumental for prevention of severe cases of COVID-19 with over 7 billion doses administered worldwide, showing exceptional safety (Ella et al., 2021; Fadlyana et al., 2021).

We developed a β-propiolactone-inactivated whole virion vaccine CoviVac which has previously shown no signs of acute/chronic, reproductive, embryo- and fetotoxicity, or teratogenic effects in the antenatal and postnatal periods of development, as well as no allergenic properties in rodents and nonhuman primates in preclinical trials (Kozlovskaya et al., 2021). Here, we report safety and immunogenicity of CoviVac in a multicenter, randomized, double-blind, placebo-controlled phase I/II clinical trial.

## MATERIALS AND METHODS

### Ethical approval

The study was conducted in accordance with the Declaration of Helsinki guidelines, ICH GCP and Russian regulations. The study protocol (VKI-I/II-08/20) and its supplementary documentation were approved by the Ethics Committee of the Ministry of Health of the Russian Federation (No. 502 from September 21, 2020). Additionally, the protocol was approved by the local Ethics Committees of the clinical sites, namely Kirov State Medical University of the Ministry of Health of Russia [No. 13/2020 from September 28, 2020]; Healthcare Unit No. 163 of FMBA of Russia, [No.1 from October 11, 2020]; Eco-Safety Scientific Research Center [No. 157 from October 1, 2020]. The study protocol was registered at clinicaltrials.gov (ID NCT05046548).

### Study design and participants

The study is a randomized, double-blind, placebo-controlled, multi-center clinical trial of the tolerability, safety, and immunogenicity (Clinical trials, phase I/II) of the inactivated whole virion concentrated purified coronavirus vaccine CoviVac in adult volunteers aged 18-60.

Safety and immunogenicity assessment was performed in 3 consequent Stages: Stage 1 included the first 15 volunteers (10 inoculated with the vaccine and 5 with placebo with 14 days interval); Stage 2 included 185 participants (140 inoculated with the vaccine and 45 with placebo) and started after the post-vaccination observation period in Stage 1; Stage 3 included 200 participants (150 inoculated with the vaccine and 50 with placebo) (Figure 1). Safety assessment was performed in Stages 1-3 and immunogenicity assessment was performed in Stage 3.

**Figure 1.**
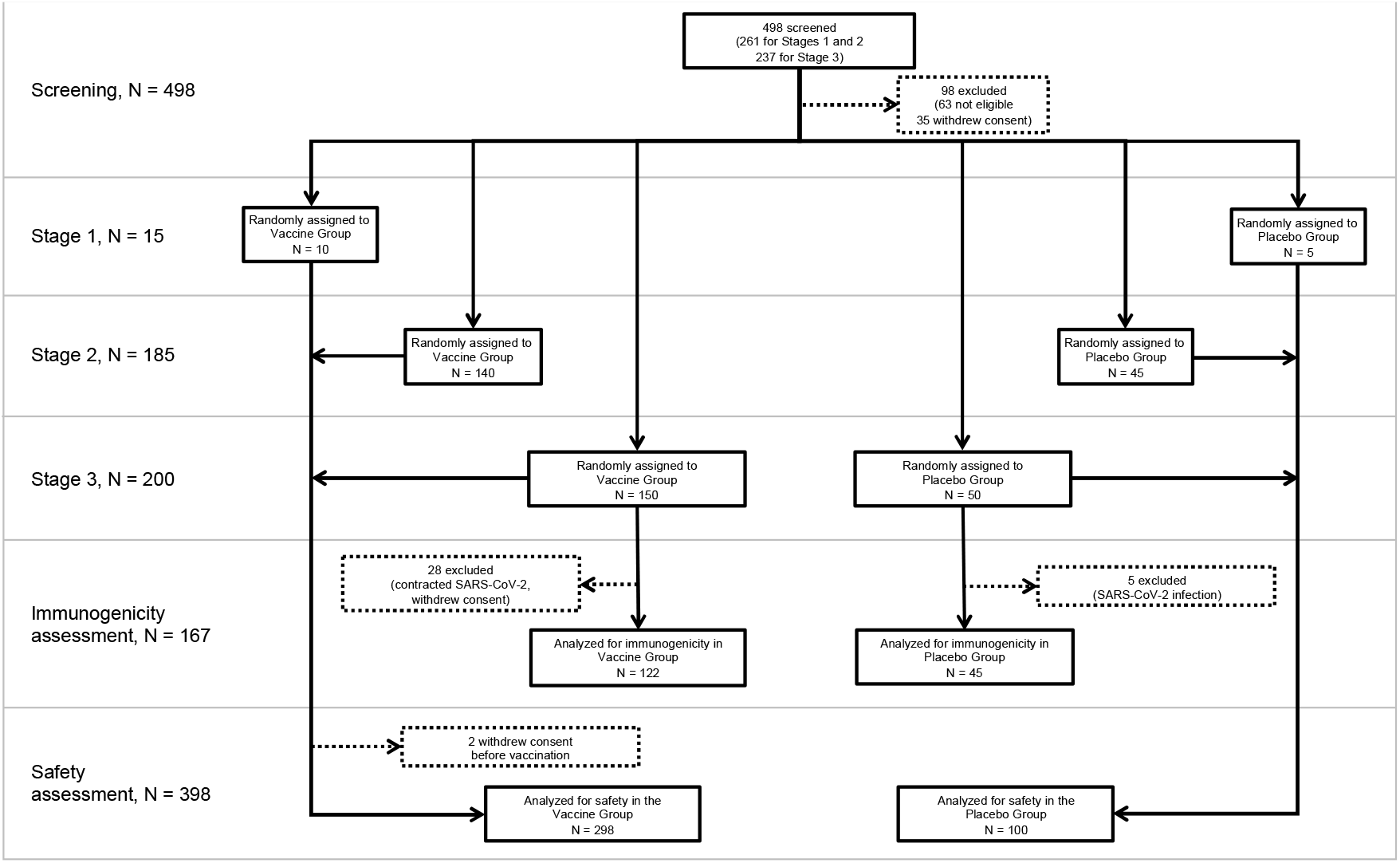
Study profile. Horizontal light grey lines represent the Stages of the study. Dotted lines represent exclusion of the participants from a given Stage of the study.

The study included participants from 18 to 60 years of age at the time of enrolment. During screening all participants were tested for SARS-CoV-2 infection by PCR in nasopharyngeal swabs. The participants of Stages 1 and 2 were also screened at enrolment for anti-SARS-CoV-2 IgM and IgG antibodies by ELISA. Individuals positive in either test were not included. The participants of Stage 3 were included regardless of their SARS-CoV-2 IgM and IgG status at enrolment. Persons with history of SARS-CoV-1, SARS-CoV-2, or MERS infection, confirmed contact with COVID-19 patients, previous severe allergic reactions, tuberculosis, cancer, mental or autoimmune diseases were also not included. All participants were screened for eligibility on the basis of their health status, including their medical history, vital signs, physical examination and laboratory test results, and were enrolled after providing signed and dated informed consent forms. Details of the inclusion, non-inclusion and exclusion criteria can be found in the study protocol (clinicaltrials.gov ID NCT05046548).

### Randomization and masking

Randomization was performed using the sealed envelope method at the day of the 1^st^ immunization. Distribution of the participants into the Vaccine Group or the Placebo Group was carried out using random number generator software.

After assigning a three-digit randomization number to a participant, a sealed envelope with the randomization number was given to a healthcare worker who administered the vaccine. The participants did not know which preparation they were receiving. After the vaccination, a separate physician observed the participant for possible adverse events (AE). The laboratory specialists were provided with the samples marked with coded numbers, and thus were also blinded.

### Procedures

CoviVac vaccine (manufactured by the Chumakov FSC R&D IBP RAS as described previously (Kozlovskaya et al., 2021)) is a whole virion β-propiolactone inactivated concentrated purified vaccine prepared in Vero cells and supplemented with aluminum hydroxide. Placebo preparation contained buffer saline used in the vaccine preparation and the equivalent amount of aluminum hydroxide.

Preparations were provided as single dose (0.5 ml) vials with sterile liquid that was injected intramuscularly into deltoid muscle in a two-dose schedule with 14 days interval. Participants were observed for any immediate AEs for 30□ min following the administration of each dose, and then daily until Day 15-17 (1-3 days after the 2^nd^ vaccination), with subsequent follow-up visits at Days 20 (only for Stages 1 and 2), 27-30 and 42-45 (for all Stages). AEs were self-recorded by participants in diaries. During the visits, vital signs measurement and physical examination were performed. Neurological assessment was performed before each vaccination, 3 days after each vaccination and 28 days after the 2^nd^ vaccination. Urine and blood samples were collected at screening and days 3, 7, 10, 14, 17, 20, 27 and 42 after the 1^st^ vaccination for hematological and biochemical analysis and immunological tests (at Stage 3). ECG was carried out at screening and once at day 2-6 after the 1^st^ vaccination.

### Safety assessment

AEs were monitored until Day 42-45 after the 1^st^ vaccination (Day 28-31 after the 2^nd^ vaccination). The solicited local AEs were pain, indurations, hematomas, swelling, itching and hypersensitivity at the injection site, and solicited systemic AEs were fever, fatigue or malaise, nervous system disorders (headache, lightheadedness), musculoskeletal and connective tissue disorders (arthralgia, myalgia), disorders of respiratory system and mediastinal organs (pain/sore throat, nasal congestion, rhinorrhea, cough, shortness of breath), gastrointestinal tract disorders (nausea, vomiting, diarrhea, impaired appetite). All unsolicited AEs were reported by participants throughout the study.

AEs were graded as mild, moderate or severe according to the severity scoring chart (Supplementary Table 1) and assessed for the probability of their relation to the tested vaccine (highly probable/certain, probable, possible, unlikely, unrelated, unknown).

### Immunogenicity assessment

Humoral immunity against SARS-CoV-2 was assessed in serum samples using chemiluminescent microparticle immunoassays (CMIA) and virus neutralization test (NT).

IgG antibodies to nucleocapsid protein (N) were detected using Architect SARS-CoV-2 IgG CMIA (Abbott Diagnostics, USA). IgG antibodies to receptor-binding domain (RBD) of the spike protein (S) were detected using Access SARS-CoV-2 IgG CMIA (Beckman, USA).

Serum samples from randomly selected participants were additionally tested in ELISA for total antibodies against RBD with CoronaPass Total (Genetico, Russia) and for total antibodies to S protein trimer (Vector-Best, Russia).

Neutralization test was performed using SARS-CoV-2 strain PIK35 in Vero cells as described previously (Kozlovskaya et al., 2021). Serum samples that did not show neutralization in 1:8 dilution were considered negative. Immunogenicity assessment was performed in 167 participants of Stage 3. Interferon-gamma (IFN-γ) production in response to stimulation with SARS-CoV-2 S-protein peptides was assessed using QuantiFERON SARS-CoV-2 Starter Set (Qiagen, Germany), using two SARS-CoV-2 S protein peptide pools (Ag1 and Ag2) designed to stimulate the production of IFN-γ by S-protein-specific CD4 (Ag1) and CD4/CD8 (Ag2) T cells, which was detected by ELISA according to the manufacturer’s protocol. A Mitogen tube served as positive control. The baseline IFN-γ level in the Nil tube (unstimulated lymphocytes) was subtracted from the IFN-γ level in the antigen tubes and the Mitogen tube to correct for background or non-specific IFN-γ signal. The results were reported in international units of IFN-γ per milliliter of whole blood (IU/mL) using a dilution curve of the standard supplied with the kit. Calibration curve was generated at each assay run. Samples were considered reactive for an IFN-γ response if the IFN-γ levels obtained from the tubes with stimulus were 0.01 IU/mL above baseline levels obtained from the unstimulated control.

### Outcomes

The primary endpoints for safety assessment were the frequency and severity of AEs within the observation period (72 h, 7 days after each vaccination and 28 days after the 2^nd^ vaccination). The primary immunogenicity endpoints were the geometric mean nAB titer (GMT) and seroconversion rate (the percentage of volunteers with 4-fold increase in GMT) 28 days after the 2^nd^ vaccination. Secondary endpoints included seroconversion rates at Days 7 and 14 and Months 2, 3, 4, 5 and 6 after the 2^nd^ vaccination.

### Statistical analysis

The sample size was determined based on the requirements of the national guidelines, and the distribution of the participants between Vaccine Group and Placebo Group was 3:1. All parameters were evaluated using descriptive statistics (mean, standard deviation, percentile, medians, min and max). All efficacy parameters are presented as mean with 95% confidence interval (CI). Value comparisons (for example, GMT) were made using ANOVA or Mann-Whitney tests. P values below 0.05 (two-sided) were considered to be significant. Frequencies (%) were compared with Chi-square test or Fisher’s exact test. Analysis of variance or Friedman’s analysis were used for comparisons of the parameters over time. Spearman’s rank correlation coefficient was used for correlation analysis.

## RESULTS

### Demographic and anthropometric data

Study profile is presented in Figure 1. Overall, starting from October 3, 2020, 498 volunteers were screened for eligibility at three clinical sites. Based on clinical, laboratory and instrumental examinations, 400 volunteers aged 18-60 years were enrolled in the study. All enrolled participants were considered healthy and had no previous or current diagnosed comorbid conditions. All participants were randomized by the sealed envelope method into 2 groups: 300 participants were assigned to receive the vaccine and 100 participants were assigned to receive placebo (2 injections, 0.5 ml, 14 days interval for both the vaccine and placebo).

Two out of 300 participants from the Vaccine Group withdrew consent prior to administration of the 1^st^ dose. Thus, safety assessment was performed in 398 (298 in the Vaccine Group and 100 in the Placebo Group) participants of the study. Participants who contracted SARS-CoV-2 infection during the observation period were not excluded from safety assessment.

Out of 200 (150 in the Vaccine Group and 50 in the Placebo Group) participants initially included into Stage 3 for immunogenicity assessment, a total of 33 (28 from the Vaccine Group and 5 from the Placebo Group) were excluded during the observation period as they contracted SARS-CoV-2 infection or withdrew consent. Thus, immunogenicity assessment was performed in 167 (122 in the Vaccine Group and 45 in the Placebo Group) participants of Stage 3.

The demographic characteristics of the participants in the safety and immunogenicity assessment populations were similar across treatment groups in terms of sex, age, and BMI (Table 1). The mean age of the study participants was 33.1 and 33.5 years, the female/male proportion was 36.9/63.1 and 34/66, and mean body mass index (BMI) was 23.8 and 24 kg/m^2^ for the Vaccine Group and the Placebo Group, correspondingly.

### Tolerance, reactogenicity and safety

In this trial, participants were monitored for AEs until Day 42-45 after the 1^st^ vaccination (Day 28-31 after the 2^nd^ vaccination). The studied vaccine has shown good tolerability and safety. No deaths, serious AEs, or other significant AEs related to vaccination have been reported. All observed severe adverse events (SAEs) were classified as unlikely to be related or unrelated to the tested vaccine.

In total, during the study period, 244 AEs were recorded in 136 (34.2 %) participants, of which 189 AEs were observed in 105 (35.2%) of Vaccine Group participants, and 55 AEs in 31 (31%) of Placebo Group participants (Table 2, Supplementary Table S2, Supplementary Table S3).

**Table 1.**
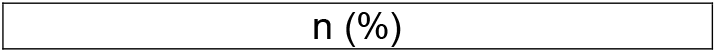
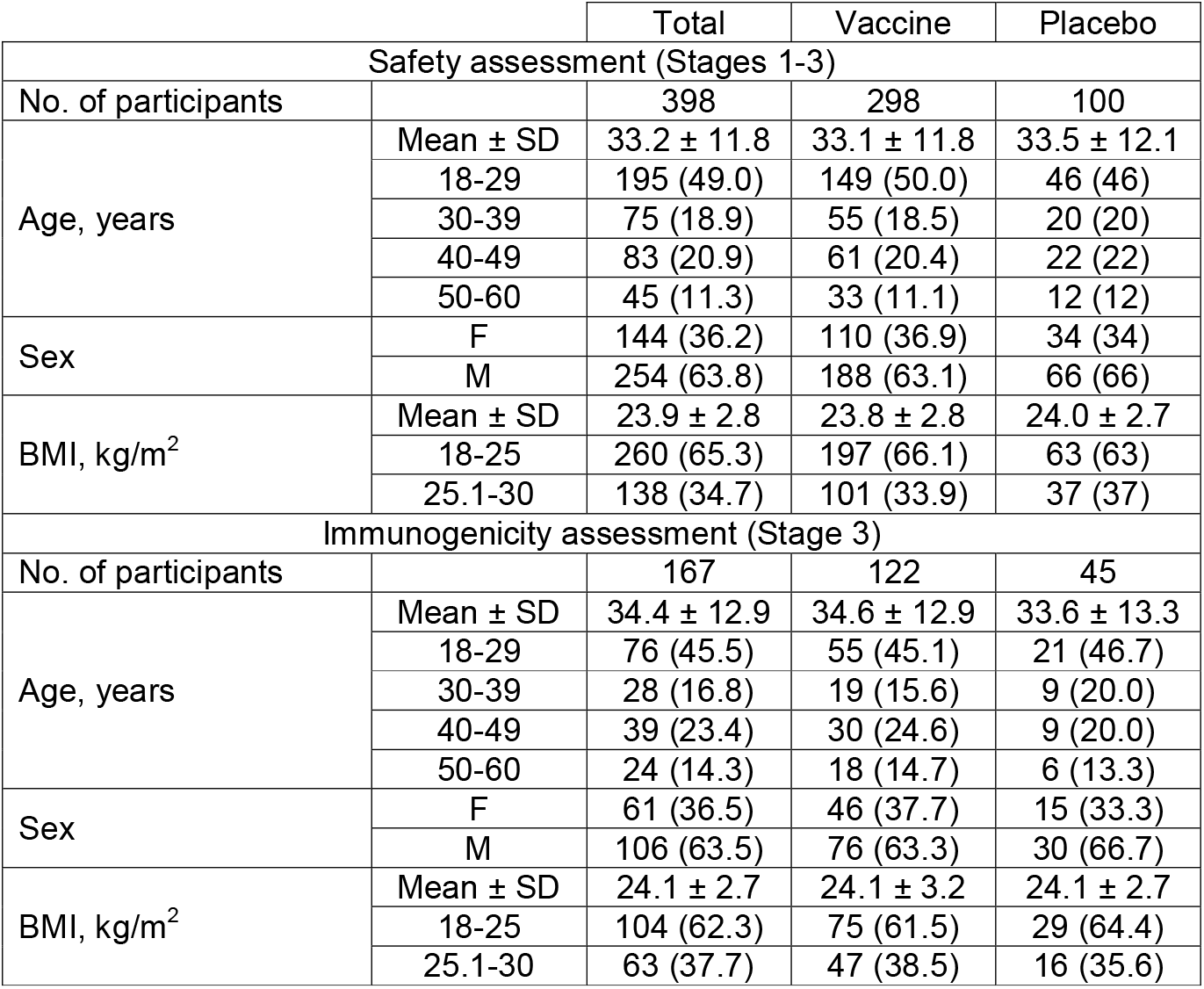
Demographic and anthropometric characteristics of the participants of the clinical trial

|  |  | Total | Vaccine | Placebo |
| --- | --- | --- | --- | --- |
| Safety assessment (Stages 1-3) |  |  |  |  |
| No. of participants |  | 398 | 298 | 100 |
| Age, years | Mean ± SD | 33.2 ± 11.8 | 33.1 ± 11.8 | 33.5 ± 12.1 |
|  | 18-29 | 195 (49.0) | 149 (50.0) | 46 (46) |
|  | 30-39 | 75 (18.9) | 55 (18.5) | 20 (20) |
|  | 40-49 | 83 (20.9) | 61 (20.4) | 22 (22) |
|  | 50-60 | 45 (11.3) | 33 (11.1) | 12 (12) |
| Sex | F | 144 (36.2) | 110 (36.9) | 34 (34) |
|  | M | 254 (63.8) | 188 (63.1) | 66 (66) |
| BMI, kg/m<sup>2</sup> | Mean ± SD | 23.9 ± 2.8 | 23.8 ± 2.8 | 24.0 ± 2.7 |
|  | 18-25 | 260 (65.3) | 197 (66.1) | 63 (63) |
|  | 25.1-30 | 138 (34.7) | 101 (33.9) | 37 (37) |
| Immunogenicity assessment (Stage 3) |  |  |  |  |
| No. of participants |  | 167 | 122 | 45 |
| Age, years | Mean ± SD | 34.4 ± 12.9 | 34.6 ± 12.9 | 33.6 ± 13.3 |
|  | 18-29 | 76 (45.5) | 55 (45.1) | 21 (46.7) |
|  | 30-39 | 28 (16.8) | 19 (15.6) | 9 (20.0) |
|  | 40-49 | 39 (23.4) | 30 (24.6) | 9 (20.0) |
|  | 50-60 | 24 (14.3) | 18 (14.7) | 6 (13.3) |
| Sex | F | 61 (36.5) | 46 (37.7) | 15 (33.3) |
|  | M | 106 (63.5) | 76 (63.3) | 30 (66.7) |
| BMI, kg/m<sup>2</sup> | Mean ± SD | 24.1 ± 2.7 | 24.1 ± 3.2 | 24.1 ± 2.7 |
|  | 18-25 | 104 (62.3) | 75 (61.5) | 29 (64.4) |
|  | 25.1-30 | 63 (37.7) | 47 (38.5) | 16 (35.6) |

**Table 2.**
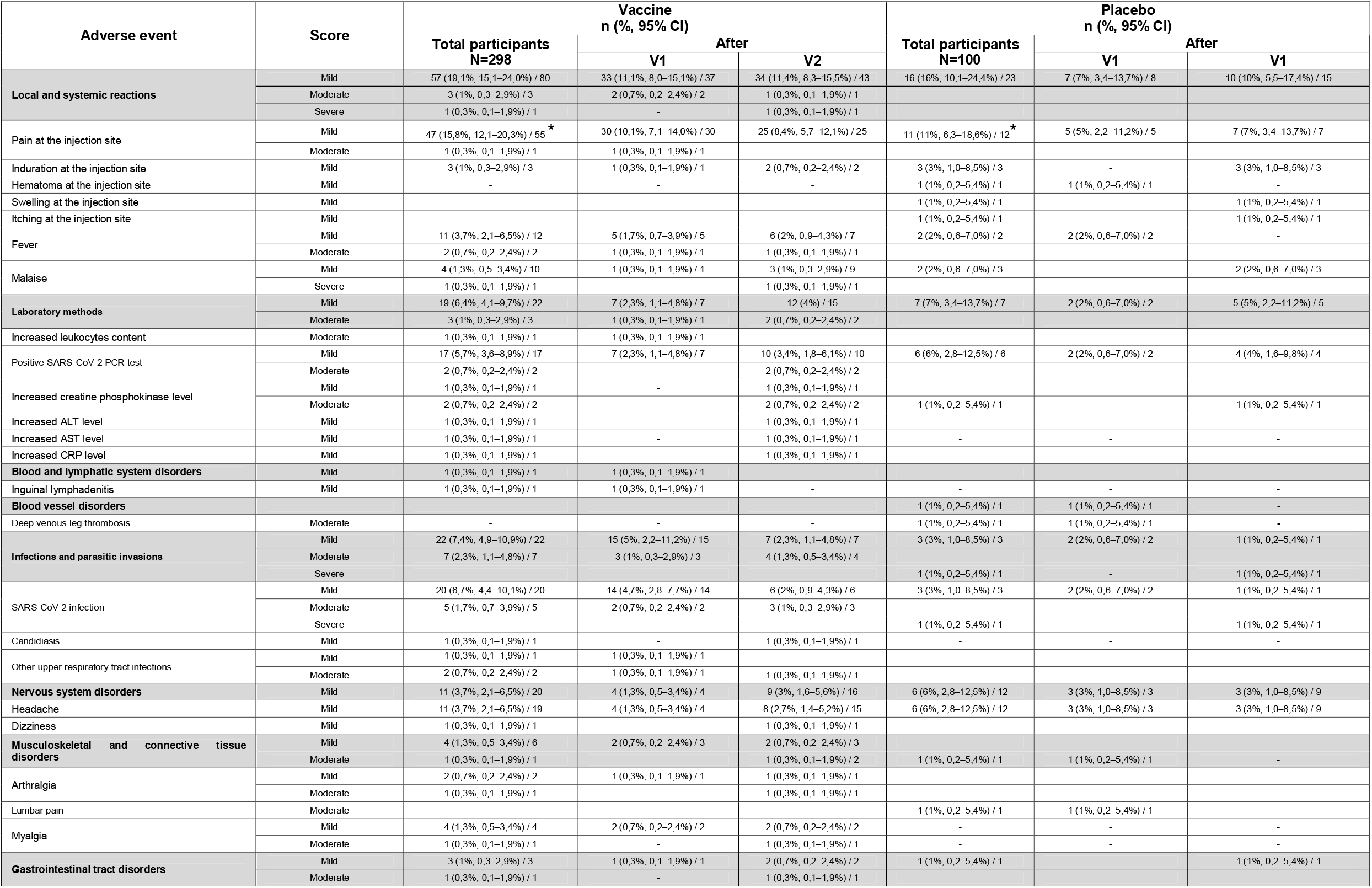
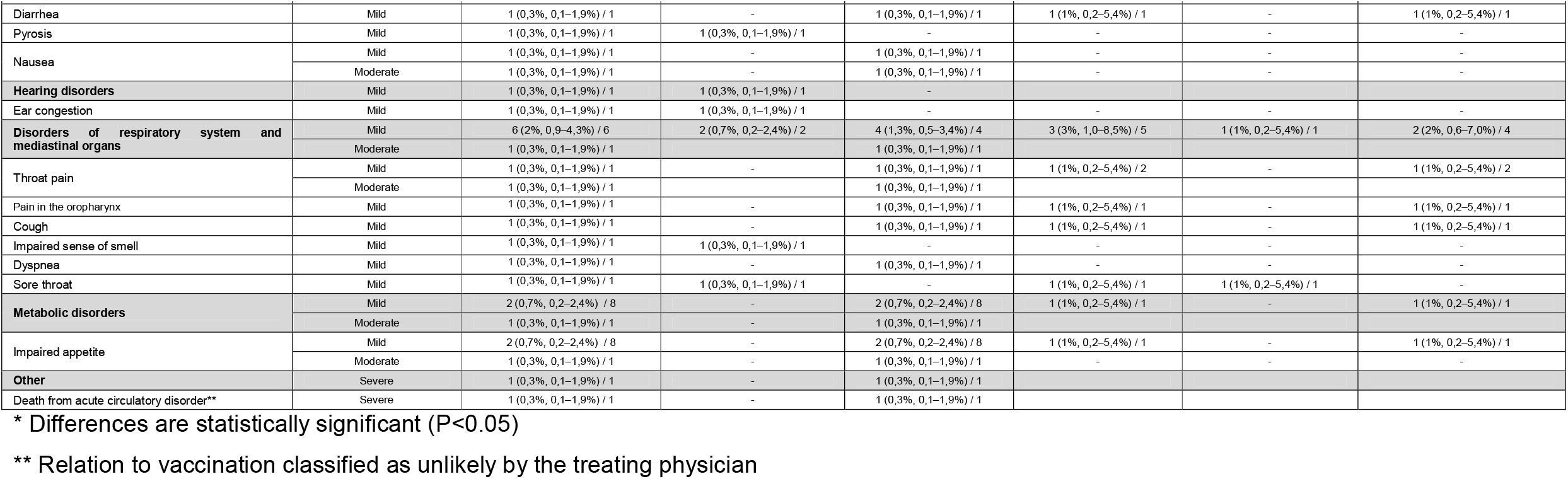
Severity of adverse events observed in the Vaccine Group and in the Placebo Group within 28 days following each vaccination

**Table 4.** Percent of participants with detectable anti-SARS-CoV-2 antibodies in NT and CMIA at different time points after the first vaccination

| Group | N | Test | Positive (% , 95% CI) |  |  |  |  |  |
| --- | --- | --- | --- | --- | --- | --- | --- | --- |
|  |  |  | Screening | Day 7 | Day 14 | Day 21<br>(7 days after<br>the 2<sup>nd</sup><br>vaccination) | Day 28<br>(14 days after<br>the 2<sup>nd</sup><br>vaccination) | Day 42<br>(28 days after<br>the 2<sup>nd</sup><br>vaccination) |
| Vaccine | 122 | nAB (NT) | 53<br>(43.4, 35.0–52.3) | 68<br>(55.7, 46.9–64.2) | 92<br>(75.4, 67.1–82.2) | 112<br>(91.8, 85.6–95.5) | 112<br>(91.8, 85.6–95.5) | 113<br>(92.6, 86.6–96.1) |
|  | 101 | anti-RBD IgG<br>(Beckman) | 28<br>(28, 20.1–37.5)<sup>1</sup> | 33<br>(32.7, 24.3–42.3) | 43<br>(42.6, 33.4–52.3) | 44<br>(44.9, 35.4–54.8)<sup>2</sup> | 47<br>(46.5, 37.1–56.2) | 44<br>(43.6, 34.3–53.3) |
|  | 101 | anti-N IgG<br>(Abbot) | 24<br>(24, 16.7–33.2)<sup>1</sup> | 28<br>(27.7, 19.9–37.1) | 34<br>(33.7, 25.2–43.3) | 43<br>(43.9, 34.5–53.7)<sup>2</sup> | 57<br>(56.4, 46.7–65.7) | 56<br>(55.4, 45.7–64.8) |
| Placebo | 45 | nAB (NT) | 17<br>(37.8, 25.1–52.4) | 18<br>(40.0, 27.0–54.6) | 19<br>(42.2, 29.0–56.7) | 17<br>(38.6, 25.7–53.4)<sup>3</sup> | 17<br>(37.8, 25.1–52.4) | 18<br>(40.9, 27.7–55.6)<sup>3</sup> |
|  | 36 | anti-RBD IgG<br>(Beckman) | 10<br>(27.8, 15.8–44) | 11<br>(30.6, 18–46.9) | 11<br>(30.6, 18–46.9) | 11<br>(30.6, 18–46.9) | 11<br>(30.6, 18–46.9) | 9<br>(25, 13.8–41.1) |
|  | 36 | anti-N IgG<br>(Abbot) | 10<br>(27.8, 15.8–44) | 10<br>(27.8, 15.8–44) | 8<br>(22.2, 11.7–38.1) | 9<br>(25, 13.8–41.1) | 8<br>(22.2, 11.7–38.1) | 8<br>(22.2, 11.7–38.1) |

<sup>1</sup> N = 100; <sup>2</sup> N = 98; <sup>3</sup> N = 44

The most common AE in Vaccine Group was pain at the injection site. It was registered in the Placebo Group as well, but was significantly less common (P<0.05).

Changes in the parameters of clinical blood tests, biochemical blood tests and general urine analysis, observed at different time points, in most cases were regarded as clinically insignificant and unrelated to vaccination (Supplementary Table S2, Supplementary Table S3). Clinically significant deviations in the parameters of creatine phosphokinase, ALT, AST, CRP and leukocytes were observed in both groups and their frequencies did not differ between the study groups. No clinically significant deviations in the parameters evaluated during physical examination, neurological status and ECG were observed in both groups.

One participant (0.3%, 0.1–1.9 95% CI) from the Vaccine Group died at Day 29 of the observation period as a result of acute circulatory disorder. Autopsy examination revealed signs of chronic mild hepatitis, pre-existing stromal cardiosclerosis and dilated cardiomyopathy manifested by myocardial hypertrophy. The character of the lesions observed at histological examination indicated long-lasting pathology of the myocardium. As the participant had no clinically significant deviations in any of the clinical parameters throughout the observation period, including physical condition, body temperature, vital signs, urine analysis, biochemical parameters of blood, complete blood count, coagulation parameters and ECG, and had no local or systemic reactions after each immunization, the relation of this SAE to vaccination was classified by the treating physician as unlikely.

### Selection of the method of humoral immunity assessment

To determine the optimal method of anti-SARS-CoV-2 humoral immunogenicity assessment we compared the neutralizing antibody (nAB) detection rates in NT with two commercial CMIA tests commonly used for the screening of anti-N protein IgG (Architect SARS-CoV-2 IgG, Abbott Diagnostics, USA) and anti-RBD IgG (Access SARS-CoV-2 IgG, Beckman, USA). We used NT and both CMIA tests to analyze serum samples from all 167 participants (122 from the Vaccine Group and 45 from the Placebo Group) included in Stage 3, obtained at screening and at Days 7, 14, 21, 28 and 42 after the 1^st^ immunization (Table 3). The analysis included both the participants who were anti-SARS-CoV-2 nAB negative (69/122 in Vaccine Group and 28/45 in Placebo Group) and anti-SARS-CoV-2 nAB positive (53/122 in Vaccine Group and 17/45 in Placebo Group) at screening. There was no significant correlation between seropositivity rates assessed by NT and CMIA at all time points (Table 3).

Therefore, further in order to assess the sensitivity of commercial CMIA and ELISA tests, serum samples from 27 randomly selected participants from the Vaccine Group in Stage 3 who were anti-SARS-CoV-2 nAB negative at screening were used to evaluate the correlation between the virus neutralization titers in NT and the positivity coefficients in binding antibody detection assays. Two abovementioned CMIA tests and two ELISA-based tests detecting total antibodies to S trimer expressed in eukaryotic system and total antibodies to RBD (SARS-CoV-2-AB total-EIA-BEST, Vector-Best, Russia and CoronaPass Total kit, Genetico, Russia, respectively) were used (Figure 2). Both ELISA-based total antibody detection kits showed higher correlation of seropositivity rates and positivity coefficients with the virus neutralization titers in NT than the CMIA tests. The Vector-Best ELISA kit was able to detect total antibodies to S trimer in 3/27 participants at Day 14 after the 1^st^ immunization (Figure 2B), while virus-neutralizing antibodies in NT (Figure 2A) and total antibodies to RBD (Figure 2C) were detected only from Day 21.

**Figure 2.**
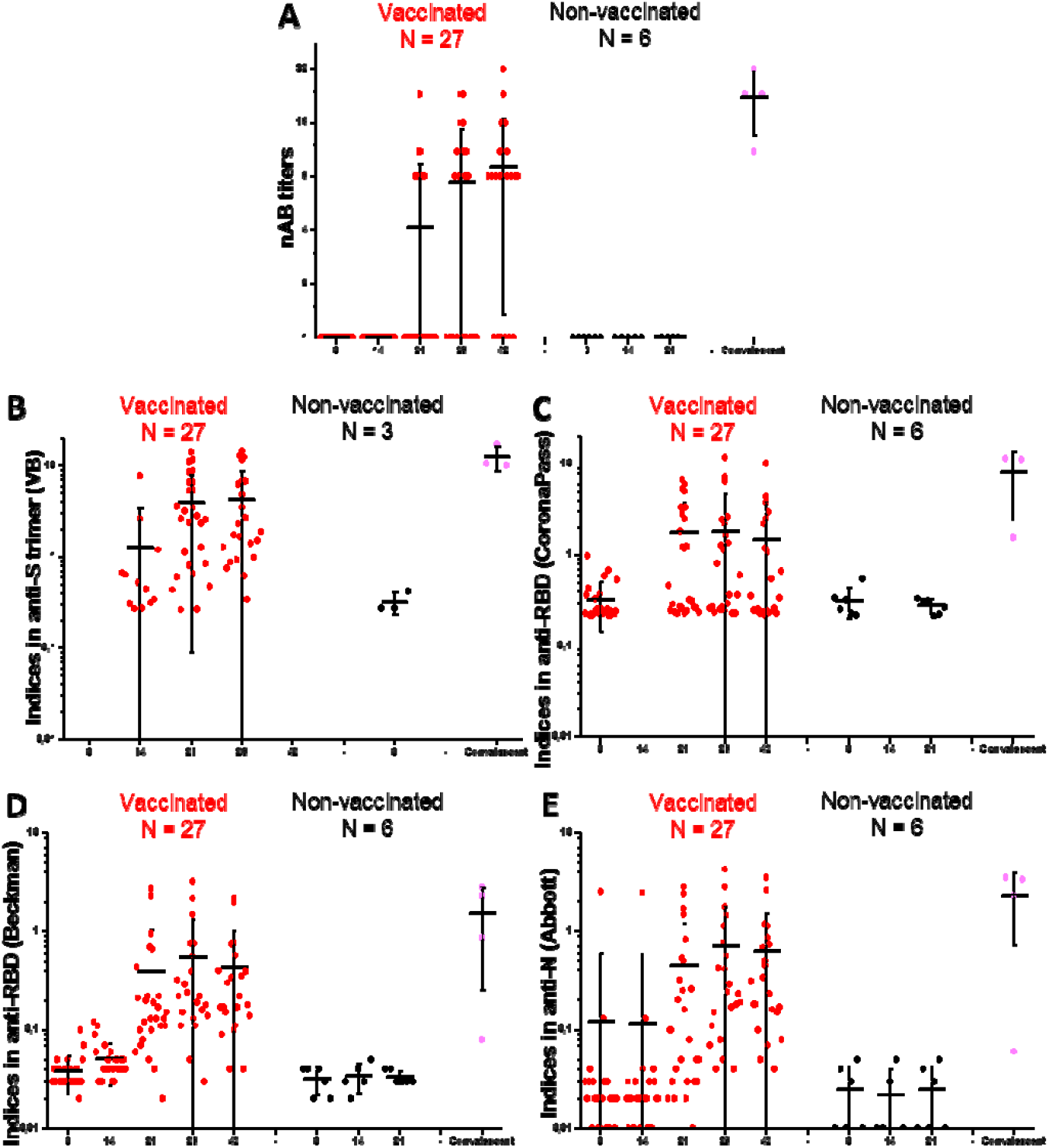
Anti-SARS-CoV-2 antibody levels following immunization of 27 randomly selected participants who were seronegative at screening. nAB titers in NT (A); and positivity coefficients for ELISA and CMIA kits: anti-S trimer total antibodies ELISA (Vector-Best) (B); anti-RBD total antibodies ELISA (CoronaPass) (C); anti-RBD IgG CMIA (Beckman) (D); and anti-N IgG CMIA (Abbot) (E). Black line – Mean, whiskers – SD. Blue area signifies a positive threshold (grey zone) for each test. Black dots represent the results from sera samples from 6 randomly selected participants from Placebo Group obtained in the same tests in parallel (with Vector-Best sera from 3 participants were tested). Purple dots represent 4 samples of convalescent sera used as positive controls.

The analysis of the individual test results showed that in the samples with detectable nAB levels we could see a statistically significant increase in the positivity coefficients in CMIA kits compared with the samples obtained at screening (Figure 3 A,D,E), but the levels were below the “grey zone”, indicating insufficient sensitivity of these tests. Thus, we did not find a strong correlation between the results of NT and the results of CMIA and ELISA tests (Figure 3E), and only NT results were used for further calculations of seroconversion rates.

**Figure 3.**
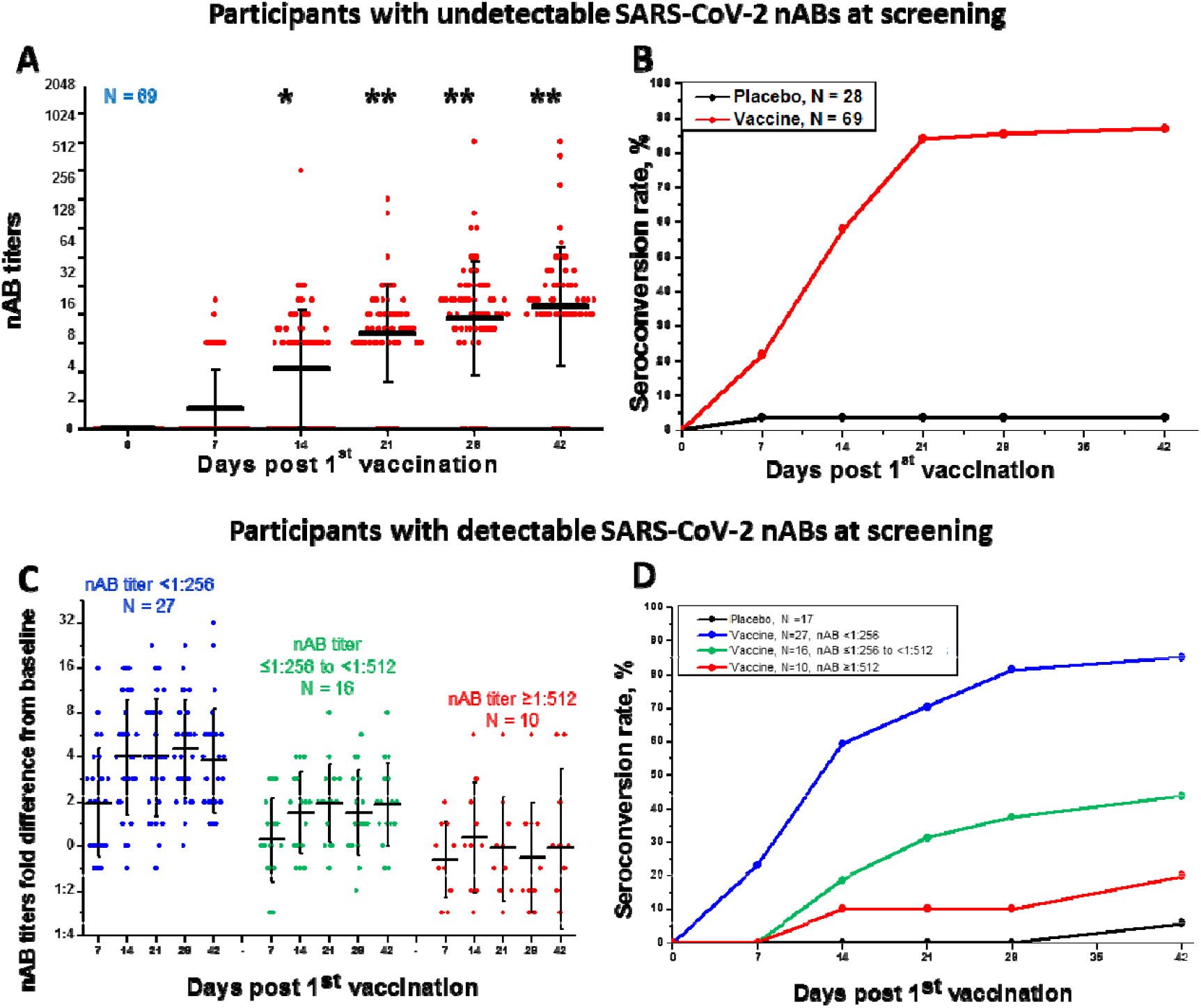
Seroconversion rates and SARS-CoV-2 neutralizing antibody (nAB) levels in sera of study participants at different time points post vaccination. SARS-CoV-2 nAB titers (A) and cumulative seroconversion rate (B) in Vaccine Group and Placebo Group participants who were seronegative at screening. SARS-CoV-2 nAB titers fold difference from baseline level (C) and cumulative seroconversion rate (defined as 4-fold increase in the nAB levels) (D) in Vaccine Group and Placebo Group participants who had detectable nABs at screening at titers <1:256 (N=27), ≤1:256-<1:512 (N=16) or ≥1:512 (N=10). Black line – Mean, whiskers – SD, * – titers at Days 14, 21, 28 and 42 significantly differ from baseline and from Day 7 [p<0.05], ** – the difference between titers at Days 14, 21, 28 and 42 is insignificant [p>0.05].

### CoviVac humoral immunogenicity assessment (NT)

Immunogenicity assessment was separately performed for the participants who were anti-SARS-CoV-2 nAB negative (69/122 in Vaccine Group and 28/45 in Placebo Group) or anti-SARS-CoV-2 nAB positive (53/122 in Vaccine Group and 17/45 in Placebo Group) at screening.

Participants with undetectable anti-SARS-CoV-2 antibodies at screening (with any method) showed increasing nAB titers and seroconversion from day 7 post 1^st^ vaccination (Figure 3A). By Day 7 after the 2^nd^ vaccination (21 days total) the seroconversion rate reached 84.1% and did not decrease during the period of observation (until Day 28 post 2^nd^ vaccination, Figure 3B). GMT of nABs peaked at Day 21 after the 1^st^ vaccination and did not decrease significantly during the period of observation (Figure 3A). Among the participants, who did not have detectable anti-SARS-CoV-2 nAB antibodies at screening, nABs were detected in 0/69 (0%), 15/69 (21.7%), 40/69 (57.9%), 58/69 (84.1%), 59/69 (85.5%) and 60/69 (86.9%) at screening, Days 7, 14, 21, 28 and 42, respectively. The GMT in positive samples at Days 7, 14, 21, 28 and 42 was 1:2, 1:4, 1:10, 1:15 and 1:20, respectively (Figure 3 A, B). According to the protocol of the clinical trial, seroconversion for participants who had detectable SARS-CoV-2 nABs at screening was defined as 4-fold increase of nAB titers after vaccination compared to baseline titers.

Participants who had detectable antibodies against SARS-CoV-2 at screening showed increased nAB titers at days 7 and 14 post 1^st^ vaccination. However, after the 2^nd^ vaccination the titers did not increase significantly (Figure 3C). Moreover, vaccination of the participants with lower nAB titers (below 1:256) had a more pronounced effect of titer increase (4-fold and more), whereas we observed almost no booster effect in participants with high baseline nAB titers (1:512 and above) (Figure 3C). Nevertheless, in participants who had nAB titers below 1:256 at screening the rate of 4-fold increase of nAB levels was comparable to the seroconversion rates for participants, who were seronegative at screening (85.2% vs. 86.9%, correspondingly, Figure 3D).

### Cellular immunity assessment

Parameters of cellular immunity were determined for the same 27 randomly selected participants from the Vaccine Group in Stage 3, who were tested during the assessment of the sensitivity of commercial CMIA and ELISA tests. Cellular immune response was assessed in whole blood of the trial participants using QuantiFERON SARS-CoV-2 Starter Set (Qiagen, Germany) by stimulation of IFN-γ production by S-protein-specific CD4 and CD4/CD8 T cells using two SARS-CoV-2 S protein peptide pools (Ag1 and Ag2, respectively). The data is summarized in Figure 4.

**Figure 4.**
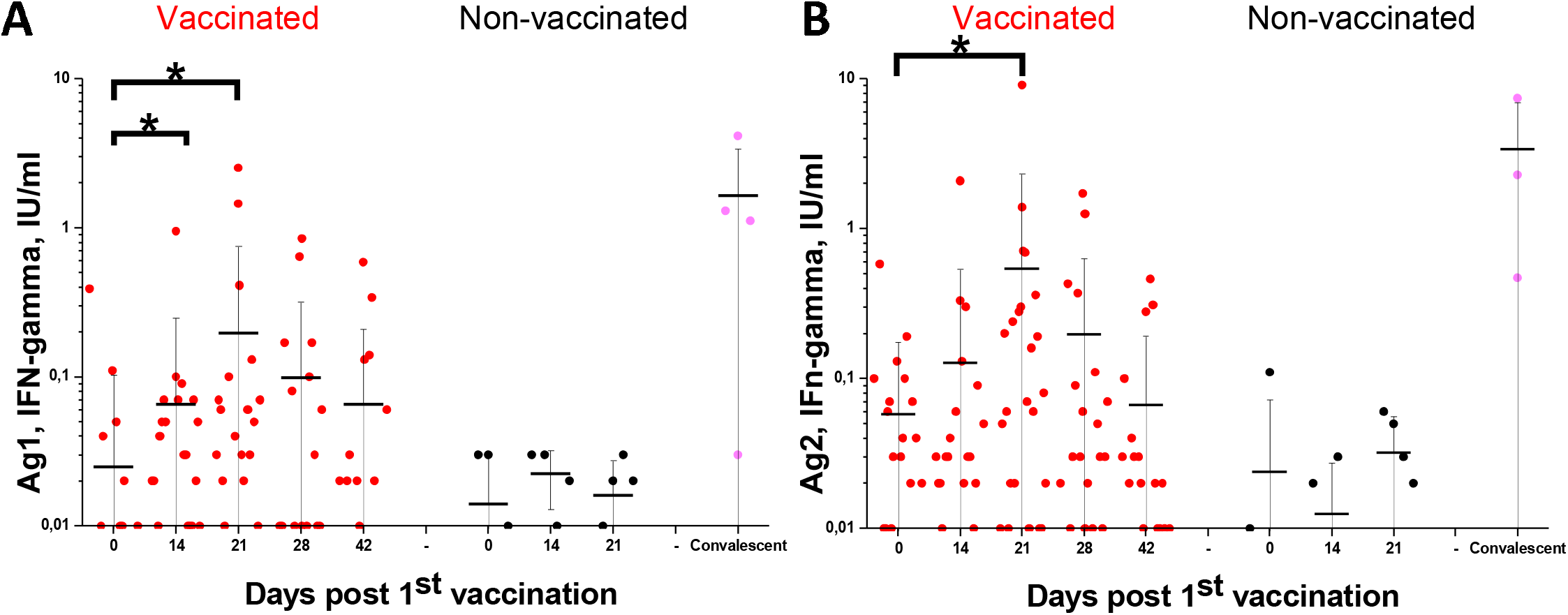
Anti-SARS-CoV-2 cellular immunity test results for randomly selected participants with undetectable SARS-CoV-2 nAB at screening at different time points post vaccination: A) IFN-gamma levels induced by QuantiFERON Ag1; B) IFN-gamma levels induced by QuantiFERON Ag2 Black line – Mean, whiskers – SD, * – difference is significant (Mann-Whitney, p<0.05).

Compared to baseline levels, we observed a statistically significant (p<0.05) increase of IFN-γ production by peptide-stimulated CD4+ cells of vaccinated participants at Days 14 and 21 after the 1^st^ immunization and a statistically significant increase of combined IFN-γ production by stimulated CD4+ and CD8+ cells at Day 21 after the 2nd immunization, indicating the development of both SARS-CoV-2 S protein-specific CD4 helper and CD8 cytotoxic cells.

## DISCUSSION

According to the WHO, by far 18 inactivated vaccines against COVID-19 have passed Phase I-II clinical trials worldwide, of which five have also passed Phase III trials (WHO, 2021). Successful clinical trials of several inactivated vaccines demonstrate that they are highly useful in prevention of severe COVID-19 cases in the situation of drastic global shortage of the COVID-19 vaccines. Previously, the Chumakov Center has successfully developed two inactivated vaccines against tick-borne encephalitis and poliomyelitis produced using Vero cells (Piniaeva et al., 2021; Vorovitch et al., 2020). Therefore, the development of the COVID-19 vaccine CoviVac was based on established cell cultivating platform using previously proven methods. Immunogenicity, safety and protective efficacy of CoviVac has been previously shown in preclinical studies (Kozlovskaya et al., 2021).

Here we present the results of a randomized, double-blind, placebo-controlled, multi-center clinical trial of the tolerability, safety, and immunogenicity of the inactivated whole virion concentrated purified coronavirus vaccine CoviVac in adult volunteers aged 18-60.

Overall, starting from October 3, 2020, 498 volunteers were screened for eligibility at three clinical sites and 400 volunteers aged 18-60 years were enrolled in the study. All participants were randomized into 2 groups: 300 participants were assigned to receive the vaccine and 100 participants were assigned to receive placebo (2 injections, 0.5 ml, 14 days interval for both the vaccine and placebo).

The studied vaccine has shown good tolerability and safety. No deaths, serious adverse events, or other significant AEs related to vaccination have been reported. The most common AE in the Vaccine Group was pain at the injection site. It was registered in the Placebo Group as well, but was significantly less common (P<0.05). One participant (0.3%, 0.1–1.9 95% CI) from the Vaccine Group died at Day 29 of the observation period as a result of acute circulatory disorder, which was not related to the tested vaccine. Autopsy examination revealed signs of chronic mild hepatitis, pre-existing stromal cardiosclerosis and dilated cardiomyopathy manifested by myocardial hypertrophy, which were undetectable by the methods used at pre-vaccination screening.

For immunogenicity screening we first assessed the sensitivity of two CMIA used for the detection of anti-SARS-CoV-2 IgG to N and S proteins and two ELISA kits widely used in the diagnostics of SARS-CoV-2 infection, to estimate if their sensitivity is sufficient to detect the post-vaccination antibodies. There was no significant correlation between seropositivity rates assessed by neutralization test (NT) and CMIA (Table 3), although we could see a statistically significant increase in the positivity indices in vaccinated participants who were seronegative at screening. This observation could have several explanations: (1) as the vaccine contains S trimers, nABs can form against other parts of S, not only RBD, (for example, to neutralizing antibody binding domain, 1-319 aa), or to nonlinear antigen sites, so, anti-RBD test systems (Beckman) cannot detect them; (2) the vaccine contains N protein, but in lower quantities than is produced during SARS-CoV-2 infection, so anti-N test systems (Abbott) cannot detect them; (3) both CMIA tests measure IgG levels, when, technically, nABs can belong to other classes. As compared to CMIA tests, both ELISA-based antibody detection kits showed higher correlation of seropositivity rates with NT results. Based on these observations, only NT results were used for further calculations of seroconversion rates.

In total, immunogenicity assessment was performed by NT in 167 volunteers (122 in Vaccine Group and 45 in Placebo Group) separately for the participants who were anti-SARS-CoV-2 nAB negative (69/122 in Vaccine Group and 28/45 in Placebo Group) or anti-SARS-CoV-2 nAB positive (53/122 in Vaccine Group and 17/45 in Placebo Group) at screening.

At Day 42 after the 1^st^ immunization the seroconversion rate in participants who were seronegative at screening was 86.9% with average the nAB GMT of 1:20. However, these nAB levels are above the estimated levels providing protection from symptomatic SARS-CoV-2 infection (Khoury et al., 2021), further protectivity assessment must be performed in Phase III trials.

Several studies have shown that antibody responses generated both after vaccination and after SARS-CoV-2 infection can decrease significantly over time (Cho et al., 2021; Prévost et al., 2020; Wang et al., 2020), which would require boosting immunizations in the long term. In this study we show the results of CoviVac immunization in participants with pre-existing anti-SARS-CoV-2 antibodies.

In this study ‘seroconversion’ for the participants who were seropositive at screening was defined as 4-fold increase in nAB titers compared to baseline. The seroconversion rate for the participants who had NT titers in NT below 1:256 at screening was comparable to that for participants who were seronegative at screening (85.2% and 86.9%, respectively), while in the participants with nAB titers >1:256 the seroconversion rate did not exceed 45%. For the participants who were seropositive at screening the 2nd immunization did not lead to a further significant increase in NT titers. Both facts signify that there is a maximum level of antibodies against SARS-CoV-2, which human immune system can produce, and further antigen addition is useless.

The cellular immune response was assessed based on the production of IFN-γ by the CD4+ and CD8+ lymphocytes after stimulation with SARS-CoV-2 S-protein peptide pools. Compared to baseline levels, we observed a statistically significant (p<0.05) increase of IFN-γ production by peptide-stimulated CD4+ cells in vaccinated participants at Days 14 and 21 after the 1^st^ immunization and a statistically significant increase of combined IFN-γ production by stimulated CD4+ and CD8+ cells at Day 21 after the 2nd immunization.

In conclusion, inactivated vaccine CoviVac has shown good tolerability and safety, with 86.9% NT seroconversion rates in participants, who were seronegative at screening. In participants who were seropositive at screening and had nAB titers below 1:256, a single immunization lead to a 4-fold increase in nAB levels in 85.2% cases. These findings indicate that CoviVac can be successfully used both for primary immunization in two-dose regimen and for booster immunization as a single dose in individuals with reduced neutralizing antibody levels.

## Data Availability

All data produced in the present work are contained in the manuscript

## ACKNOWLEDGMENTS

We would like to thank the study participants, the staff of the Research Centres, members of the test management groups, and the staff of Research and Development department of the Chumakov FSC R&D IBP RAS and particularly Vladislav Vasilenko, Konstantin Kaa, Zhad Mazhed, Saglara Sandzhieva, Irina Tcelykh for their contribution in the development of vaccine production process; and Aleksandra Kartseva for help in organizing clinical studies.

