## Supplementary Table 1 for "Safety and immunogenicity of inactivated whole virion vaccine CoviVac against COVID-19: a multicenter, randomized, double-blind, placebo-controlled phase I/II clinical trial"

### SUPPLEMENTARY MATERIALS

**Supplementary Table 1.** Adverse events severity score

| **Score** | **Hyperemia/swelling/induration**  **at the injection site** | **Fever** | **Other** |
| --- | --- | --- | --- |
| 0 – absent | none | ≤ 37.0 °С | none |
| 1 – mild | hyperemia <50 mm or swelling/induration <25 mm | > 37.0 °С – ≤ 37.5 °С | does not interfere  with daily activities |
| 2 – moderate | hyperemia >50 mm or swelling/induration 26-50 mm | > 37.6 °С – ≤ 38.5 °С | interferes  with daily activities |
| 3 – severe | swelling/induration >50 mm | > 38.6 °С | prevents daily activities |

**Supplementary Table 2.** Total adverse events observed in the Vaccine Group and in the Placebo Group within 28 days following each vaccination

| **Adverse event** | **Vaccine**  **Number of participants with AEs (%, 95% CI), number of AEs** | | | **Placebo**  **Number of participants with AEs (%, 95% CI), number of AEs** | | |
| --- | --- | --- | --- | --- | --- | --- |
|  | **Total participants**  **N=298** | **After** | | **Total participants**  **N=100** | **After** | |
|  |  | **V1** | **V2** |  | **V1** | **V2** |
| **Local and systemic reactions** | 58 (19,5%, 15,4–24,3%) / 84 | 34 (11,4%, 8,3–15,5%) / 39 | 34 (11,4%, 8,3–15,5%) / 45 | 16 (16%, 10,1–24,4%) / 23 | 7 (7%, 3,4–13,7%) / 8 | 10 (10%) / 15 |
| Pain at the injection site | 48 (16,1%, 12,4–20,7%) / 56 | 31 (10,4%, 7,4–14,4%) / 31 | 25 (8,4%, 5,7–12,1%) / 25 | 11 (11%, 6,3–18,6%) / 12 | 5 (5%, 2,2–11,2%) / 5 | 7 (7%, 3,4–13,7%) / 7 |
| Induration at the injection site | 3 (1%, 0,3–2,9%) / 3 | 1 (0,3%, 0,1–1,9%) / 1 | 2 (0,7%, 0,2–2,4%) / 2 | 3 (3%, 1,0–8,5%) / 3 | - | 3 (3%, 1,0–8,5%) / 3 |
| Hematoma at the injection site | - | - | - | 1 (1%, 0,2–5,4%) / 1 | 1 (1%, 0,2–5,4%) / 1 | - |
| Swelling at the injection site |  |  |  | 1 (1%, 0,2–5,4%) / 1 |  | 1 (1%, 0,2–5,4%) / 1 |
| Itching at the injection site |  |  |  | 1 (1%, 0,2–5,4%) / 1 |  | 1 (1%, 0,2–5,4%) / 1 |
| Fever | 13 (4,4%, 2,7–7,3%) / 14 | 6 (2%, 0,9–4,3%) / 6 | 7 (2,3%, 1,1–4,8%) / 8 | 2 (2%, 0,6–7,0%) / 2 | 2 (2%, 0,6–7,0%) / 2 | - |
| Malaise | 5 (1,7%, 0,7–3,9%) / 11 | 1 (0,3%, 0,1–1,9%) / 1 | 4 (1,3%, 0,5–3,4%) / 10 | 2 (2%, 0,6–7,0%) / 3 | - | 2 (2%, 0,6–7,0%) / 3 |
| **Laboratory methods** | 22 (7,4%, 4,9–10,9%) / 25 | 8 (2,7%, 1,4–5,2%) / 8 | 14 (4,7%, 2,8–7,7%) / 17 | 7 (7%, 3,4–13,7%) / 7 | 2 (2%, 0,6–7,0%) / 2 | 5 (5%, 2,2–11,2%) / 5 |
| Increased leukocytes content | 1 (0,3%, 0,1–1,9%) / 1 | 1 (0,3%, 0,1–1,9%) / 1 | - | - | - | - |
| Positive SARS-CoV-2 PCR test | 18 (6%, 3,9–9,3%) / 18 | 7 (2,3%, 1,1–4,8%) / 7 | 11 (3,7%, 2,1–6,5%) / 11 | 6 (6%, 2,8–12,5%) / 6 | 2 (2%, 0,6–7,0%) / 2 | 4 (4%, 1,6–9,8%) / 4 |
| Increased creatine phosphokinase level | 3 (1%, 0,3–2,9%) / 3 | - | 3 (1%, 0,3–2,9%) / 3 | 1 (1%, 0,2–5,4%) / 1 | - | 1 (1%, 0,2–5,4%) / 1 |
| Increased ALT level | 1 (0,3%, 0,1–1,9%) / 1 | - | 1 (0,3%, 0,1–1,9%) / 1 | - | - | - |
| Increased AST level | 1 (0,3%, 0,1–1,9%) / 1 | - | 1 (0,3%, 0,1–1,9%) / 1 | - | - | - |
| Increased CRP level | 1 (0,3%, 0,1–1,9%) / 1 | - | 1 (0,3%, 0,1–1,9%) / 1 | - | - | - |
| **Blood and lymphatic system disorders** | 1 (0,3%, 0,1–1,9%) / 1 | 1 (0,3%, 0,1–1,9%) / 1 | - |  |  |  |
| Inguinal lymphadenitis | 1 (0,3%, 0,1–1,9%) / 1 | 1 (0,3%, 0,1–1,9%) / 1 | - | - | - | - |
| **Blood vessel disorders** |  |  |  | 1 (1%, 0,2–5,4%) / 1 | 1 (1%, 0,2–5,4%) / 1 |  |
| Deep venous leg thrombosis | - | - | - | 1 (1%, 0,2–5,4%) / 1 | 1 (1%, 0,2–5,4%) / 1 | - |
| **Infections and parasitic invasions** | 29 (9,7%, 6,9–13,6%) / 29 | 18 (6%, 3,9–9,3%) / 18 | 11 (3,7%, 2,1–6,5%) / 11 | 4 (4%, 1,6–9,8%) / 4 | 2 (2%, 0,6–7,0%) / 2 | 2 (2%, 0,6–7,0%) / 2 |
| SARS-CoV-2 infection | 25 (8,4%, 5,7–12,1%) / 25 | 16 (5,4%, 3,3–8,5%) / 16 | 9 (3%, 1,6–5,6%) / 9 | 4 (4%, 1,6–9,8%) / 4 | 2 (2%, 0,6–7,0%) / 2 | 2 (2%, 0,6–7,0%) / 2 |
| Other upper respiratory tract infections | 3 (1%, 0,3–2,9%) / 3 | 2 (0,7%, 0,2–2,4%) / 2 | 1 (0,3%, 0,1–1,9%) / 1 | - | - | - |
| Candidiasis | 1 (0,3%, 0,1–1,9%) / 1 | - | 1 (0,3%, 0,1–1,9%) / 1 | - | - | - |
| **Nervous system disorders** | 11 (3,7%, 2,1–6,5%) / 20 | 4 (1,3%, 0,5–3,4%) / 4 | 9 (3%, 1,6–5,6%) / 16 | 6 (6%, 2,8–12,5%) / 12 | 3 (3%, 1,0–8,5%) / 3 | 3 (3%, 1,0–8,5%) / 9 |
| Headache | 11 (3,7%, 2,1–6,5%) / 19 | 4 (1,3%, 0,5–3,4%) / 4 | 8 (2,7%, 1,4–5,2%) / 15 | 6 (6%, 2,8–12,5%) / 12 | 3 (3%, 1,0–8,5%) / 3 | 3 (3%, 1,0–8,5%) / 9 |
| Dizziness | 1 (0,3%, 0,1–1,9%) / 1 | - | 1 (0,3%, 0,1–1,9%) / 1 | - | - | - |
| **Musculoskeletal and connective tissue disorders** | 5 (1,7%, 0,7–3,9%) / 8 | 2 (0,7%, 0,2–2,4%) / 3 | 3 (1%, 0,3–2,9%) / 5 | 1 (1%, 0,2–5,4%) / 1 | 1 (1%, 0,2–5,4%) / 1 | - |
| Arthralgia | 3 (1%, 0,3–2,9%) / 3 | 1 (0,3%, 0,1–1,9%) / 1 | 2 (0,7%, 0,2–2,4%) / 2 | - | - | - |
| Lumbar pain | - | - | - | 1 (1%, 0,2–5,4%) / 1 | 1 (1%, 0,2–5,4%) / 1 | - |
| Myalgia | 5 (1,7%, 0,7–3,9%) / 5 | 2 (0,7%, 0,2–2,4%) / 2 | 3 (1%, 0,3–2,9%) / 3 | - | - | - |
| **Gastrointestinal tract disorders** | 3 (1%, 0,3–2,9%) / 4 | 1 (0,3%, 0,1–1,9%) / 1 | 2 (0,7%, 0,2–2,4%) / 3 | 1 (1%, 0,2–5,4%) / 1 | - | 1 (1%, 0,2–5,4%) / 1 |
| Диарея | 1 (0,3%, 0,1–1,9%) / 1 | - | 1 (0,3%, 0,1–1,9%) / 1 | 1 (1%, 0,2–5,4%) / 1 | - | 1 (1%, 0,2–5,4%) / 1 |
| Diarrhea | 1 (0,3%, 0,1–1,9%) / 1 | 1 (0,3%, 0,1–1,9%) / 1 | - | - | - | - |
| Pyrosis | 2 (0,7%, 0,2–2,4%) / 2 | - | 2 (0,7%, 0,2–2,4%) / 2 | - | - | - |
| **Hearing disorders** | 1 (0,3%, 0,1–1,9%) / 1 | 1 (0,3%, 0,1–1,9%) / 1 | - |  |  |  |
| Ear congestion | 1 (0,3%, 0,1–1,9%) / 1 | 1 (0,3%, 0,1–1,9%) / 1 | - | - | - | - |
| **Disorders of respiratory system and mediastinal organs** | 7 (2,3%, 1,1–4,8%) / 7 | 2 (0,7%, 0,2–2,4%) / 2 | 5 (1,7%, 0,7–3,9%) / 5 | 3 (3%, 1,0–8,5%) / 5 | 1 (1%, 0,2–5,4%) / 1 | 2 (2%, 0,6–7,0%) / 4 |
| Throat pain | 2 (0,7%, 0,2–2,4%) / 2 | - | 2 (0,7%, 0,2–2,4%) / 2 | 1 (1%, 0,2–5,4%) / 2 | - | 1 (1%, 0,2–5,4%) / 2 |
| Боль в ротоглотке | 1 (0,3%, 0,1–1,9%) / 1 | - | 1 (0,3%, 0,1–1,9%) / 1 | 1 (1%, 0,2–5,4%) / 1 | - | 1 (1%, 0,2–5,4%) / 1 |
| Pain in the oropharynx | 1 (0,3%, 0,1–1,9%) / 1 | - | 1 (0,3%, 0,1–1,9%) / 1 | 1 (1%, 0,2–5,4%) / 1 | - | 1 (1%, 0,2–5,4%) / 1 |
| Cough | 1 (0,3%, 0,1–1,9%) / 1 | 1 (0,3%, 0,1–1,9%) / 1 | - | - | - | - |
| Impaired sense of smell | 1 (0,3%, 0,1–1,9%) / 1 | - | 1 (0,3%, 0,1–1,9%) / 1 | - | - | - |
| Dyspnea | 1 (0,3%, 0,1–1,9%) / 1 | 1 (0,3%, 0,1–1,9%) / 1 | - | 1 (1%, 0,2–5,4%) / 1 | 1 (1%, 0,2–5,4%) / 1 | - |
| **Metabolic disorders** | 3 (1%, 0,3–2,9%) / 9 | - | 3 (1%, 0,3–2,9%) / 9 | 1 (1%, 0,2–5,4%) / 1 | - | 1 (1%, 0,2–5,4%) / 1 |
| Impaired appetite | 3 (1%, 0,3–2,9%) / 9 | - | 3 (1%, 0,3–2,9%) / 9 | 1 (1%, 0,2–5,4%) / 1 | - | 1 (1%, 0,2–5,4%) / 1 |
| **Other** | 1 (0,3%, 0,1–1,9%) / 1 | - | 1 (0,3%, 0,1–1,9%) / 1 |  |  |  |
| Death from acute circulatory disorder | 1 (0,3%, 0,1–1,9%) / 1 | - | 1 (0,3%, 0,1–1,9%) / 1 |  |  |  |

**Supplementary Table 3.** Adverse events observed in the Vaccine Group and in the Placebo Group within 28 days following each vaccination by relation to vaccination

| **Adverse event** | **Relation to vaccination** | **Vaccine**  **Number of participants with AEs (%, 95% CI), number of AEs** | | | **Placebo**  **Number of participants with AEs (%, 95% CI), number of AEs** | | |
| --- | --- | --- | --- | --- | --- | --- | --- |
|  |  | **Total participants**  **N=298** | **After** | | **Total participants**  **N=100** | **After** | |
|  |  |  | **V1** | **V2** |  | **V1** | **V1** |
| **Local and systemic reactions** | Definite | 47 (15,8%, 12,1–20,3%) / 58 | 31 (10,4%, 7,4–14,4%) / 32 | 25 (8,4%, 5,7–12,1%) / 26 | 11 (11%, 6,3–18,6%) / 14 | 5 (5%, 2,2–11,2%) / 6 | 7 (7%, 3,4–13,7%) / 8 |
|  | Probable | 3 (1%, 0,3–2,9%) / 3 | 1 (0,3%, 0,1–1,9%) / 1 | 2 (0,7%, 0,2–2,4%) / 2 | 1 (1%, 0,2–5,4%) / 4 | - | 1 (1%, 0,2–5,4%) / 4 |
|  | Possible | 5 (1,7%, 0,7–3,9%) / 6 | 3 (1%, 0,3–2,9%) / 4 | 2 (0,7%, 0,2–2,4%) / 2 |  |  |  |
|  | Unlikely | 3 (1%, 0,3–2,9%) / 3 |  | 3 (1%, 0,3–2,9%) / 3 | 1 (1%, 0,2–5,4%) / 1 |  | 1 (1%, 0,2–5,4%) / 1 |
|  | Unrelated | 5 (1,7%, 0,7–3,9%) / 13 | 2 (0,7%, 0,2–2,4%) / 2 | 3 (1%, 0,3–2,9%) / 11 | 2 (2%, 0,6–7,0%) / 2 | 2 (2%, 0,6–7,0%) / 2 |  |
|  | Unknown | 1 (0,3%, 0,1–1,9%) / 1 | - | 1 (0,3%, 0,1–1,9%) / 1 | 1 (1%, 0,2–5,4%) / 2 | - | 1 (1%, 0,2–5,4%) / 2 |
| Pain at the injection site | Definite | 46 (15,4%, 11,8–20,0%) / 54 | 30 (10,1%, 7,1–14,0%) / 30 | 24 (8,1%, 5,5–11,7%) / 24 | 10 (10%, 5,5–17,4%) / 11 | 5 (5%, 2,2–11,2%) / 5 | 6 (6%, 2,8–12,5%) / 6 |
|  | Probable |  |  |  | 1 (1%, 0,2–5,4%) / 1 | - | 1 (1%, 0,2–5,4%) / 1 |
|  | Possible | 2 (0,7%, 0,2–2,4%) / 2 | 1 (0,3%, 0,1–1,9%) / 1 | 1 (0,3%, 0,1–1,9%) / 1 |  |  |  |
| Induration at the injection site | Definite | 3 (1%, 0,3–2,9%) / 3 | 1 (0,3%, 0,1–1,9%) / 1 | 2 (0,7%, 0,2–2,4%) / 2 | 2 (2%, 0,6–7,0%) / 2 | - | 2 (2%, 0,6–7,0%) / 2 |
|  | Probable |  |  |  | 1 (1%, 0,2–5,4%) / 1 |  | 1 (1%, 0,2–5,4%) / 1 |
| Hematoma at the injection site | Definite | - | - | - | 1 (1%, 0,2–5,4%) / 1 | 1 (1%, 0,2–5,4%) / 1 | - |
| Swelling at the injection site | Probable |  |  |  | 1 (1%, 0,2–5,4%) / 1 |  | 1 (1%, 0,2–5,4%) / 1 |
| Itching at the injection site | Probable |  |  |  | 1 (1%, 0,2–5,4%) / 1 |  | 1 (1%, 0,2–5,4%) / 1 |
| Fever | Definite | 1 (0,3%, 0,1–1,9%) / 1 | 1 (0,3%, 0,1–1,9%) / 1 | - | - | - | - |
|  | Probable | 3 (1%, 0,3–2,9%) / 3 | 1 (0,3%, 0,1–1,9%) / 1 | 2 (0,7%, 0,2–2,4%) / 2 | - |  | - |
|  | Possible | 3 (1%, 0,3–2,9%) / 3 | 2 (0,7%, 0,2–2,4%) / 2 | 1 (0,3%, 0,1–1,9%) / 1 | - | - | - |
|  | Unlikely | 2 (0,7%, 0,2–2,4%) / 2 | - | 2 (0,7%, 0,2–2,4%) / 2 | - | - | - |
|  | Unrelated | 4 (1,3%, 0,5–3,4%) / 5 | 2 (0,7%, 0,2–2,4%) / 2 | 2 (0,7%, 0,2–2,4%) / 3 | 2 (2%, 0,6–7,0%) / 2 | 2 (2%, 0,6–7,0%) / 2 | - |
| Malaise | Possible | 1 (0,3%, 0,1–1,9%) / 1 | 1 (0,3%, 0,1–1,9%) / 1 | - | - | - | - |
|  | Unlikely | 1 (0,3%, 0,1–1,9%) / 1 |  | 1 (0,3%, 0,1–1,9%) / 1 | 1 (1%, 0,2–5,4%) / 1 |  | 1 (1%, 0,2–5,4%) / 1 |
|  | Unrelated | 2 (0,7%, 0,2–2,4%) / 8 | - | 2 (0,7%, 0,2–2,4%) / 8 | 1 (1%, 0,2–5,4%) / 2 | - | 1 (1%, 0,2–5,4%) / 2 |
|  | Unknown | 1 (0,3%, 0,1–1,9%) / 1 |  | 1 (0,3%, 0,1–1,9%) / 1 |  |  |  |
| **Laboratory methods** | Possible | 2 (0,7%, 0,2–2,4%) / 5 |  | 2 (0,7%, 0,2–2,4%) / 5 |  |  |  |
|  | Unlikely | 1 (0,3%, 0,1–1,9%) / 1 | 1 (0,3%, 0,1–1,9%) / 1 | - |  |  |  |
|  | Unrelated | 19 (6,4%, 4,1–9,7%) / 19 | 7 (2,3%, 1,1–4,8%) / 7 | 12 (4%, 2,3–6,9%) / 12 | 7 (7%, 3,4–13,7%) / 7 | 2 (2%, 0,6–7,0%) / 2 | 5 (5%, 2,2–11,2%) / 5 |
| Increased leukocytes content | Unlikely | 1 (0,3%, 0,1–1,9%) / 1 | 1 (0,3%, 0,1–1,9%) / 1 | - | - | - | - |
| Positive SARS-CoV-2 PCR test | Unrelated | 18 (6%, 3,9–9,3%) / 18 | 7 (2,3%, 1,1–4,8%) / 7 | 11 (3,7%, 2,1–6,5%) / 11 | 6 (6%, 2,8–12,5%) / 6 | 2 (2%, 0,6–7,0%) / 2 | 4 (4%, 1,6–9,8%) / 4 |
| Increased creatine phosphokinase level | Possible | 2 (0,7%, 0,2–2,4%) / 2 |  | 2 (0,7%, 0,2–2,4%) / 2 |  |  |  |
|  | Unrelated | 1 (0,3%, 0,1–1,9%) / 1 | - | 1 (0,3%, 0,1–1,9%) / 1 | 1 (1%, 0,2–5,4%) / 1 | - | 1 (1%, 0,2–5,4%) / 1 |
| Increased ALT level | Possible | 1 (0,3%, 0,1–1,9%) / 1 | - | 1 (0,3%, 0,1–1,9%) / 1 | - | - | - |
| Increased AST level | Possible | 1 (0,3%, 0,1–1,9%) / 1 | - | 1 (0,3%, 0,1–1,9%) / 1 | - | - | - |
| Increased CRP level | Possible | 1 (0,3%, 0,1–1,9%) / 1 | - | 1 (0,3%, 0,1–1,9%) / 1 | - | - | - |
| **Blood and lymphatic system disorders** | Unlikely | 1 (0,3%, 0,1–1,9%) / 1 | 1 (0,3%, 0,1–1,9%) / 1 | - |  |  |  |
| Inguinal lymphadenitis | Unlikely | 1 (0,3%, 0,1–1,9%) / 1 | 1 (0,3%, 0,1–1,9%) / 1 | - | - | - | - |
| **Blood vessel disorders** |  |  |  |  | 1 (1%, 0,2–5,4%) / 1 | 1 (1%, 0,2–5,4%) / 1 | - |
| Deep venous leg thrombosis | Unrelated | - | - | - | 1 (1%, 0,2–5,4%) / 1 | 1 (1%, 0,2–5,4%) / 1 | - |
| **Infections and parasitic invasions** | Possible | 1 (0,3%, 0,1–1,9%) / 1 | 1 (0,3%, 0,1–1,9%) / 1 | - |  |  |  |
|  | Unlikely | 1 (0,3%, 0,1–1,9%) / 1 | - | 1 (0,3%, 0,1–1,9%) / 1 |  |  |  |
|  | Unrelated | 27 (9,1%, 6,3–12,9%) / 27 | 17 (5,7%, 3,6–8,9%) / 17 | 10 (3,4%, 1,8–6,1%) / 10 | 4 (4%, 1,6–9,8%) / 4 | 2 (2%, 0,6–7,0%) / 2 | 2 (2%, 0,6–7,0%) / 2 |
| SARS-CoV-2 infection | Unrelated | 25 (8,4%, 5,7–12,1%) / 25 | 16 (5,4%, 3,3–8,5%) / 16 | 9 (3%, 1,6–5,6%) / 9 | 4 (4%, 1,6–9,8%) / 4 | 2 (2%, 0,6–7,0%) / 2 | 2 (2%, 0,6–7,0%) / 2 |
| Candidiasis | Unlikely | 1 (0,3%, 0,1–1,9%) / 1 | - | 1 (0,3%, 0,1–1,9%) / 1 | - | - | - |
| Other upper respiratory tract infections | Possible | 1 (0,3%, 0,1–1,9%) / 1 | 1 (0,3%, 0,1–1,9%) / 1 | - |  |  |  |
|  | Unrelated | 2 (0,7%, 0,2–2,4%) / 2 | 1 (0,3%, 0,1–1,9%) / 1 | 1 (0,3%, 0,1–1,9%) / 1 |  |  |  |
| **Nervous system disorders** | Possible | 6 (2%, 0,9–4,3%) / 7 | 3 (1%, 0,3–2,9%) / 3 | 4 (1,3%, 0,5–3,4%) / 4 | 1 (1%, 0,2–5,4%) / 1 | 1 (1%, 0,2–5,4%) / 1 | - |
|  | Unlikely | 2 (0,7%, 0,2–2,4%) / 3 | - | 2 (0,7%, 0,2–2,4%) / 3 | 1 (1%, 0,2–5,4%) / 1 | 1 (1%, 0,2–5,4%) / 1 | - |
|  | Unrelated | 4 (1,3%, 0,5–3,4%) / 9 |  | 4 (1,3%, 0,5–3,4%) / 9 | 3 (3%, 1,0–8,5%) / 8 | 1 (1%, 0,2–5,4%) / 1 | 2 (2%, 0,6–7,0%) / 7 |
|  | Unknown | 1 (0,3%, 0,1–1,9%) / 1 | 1 (0,3%, 0,1–1,9%) / 1 | - | 1 (1%, 0,2–5,4%) / 2 |  | 1 (1%, 0,2–5,4%) / 2 |
| Headache | Possible | 6 (2%, 0,9–4,3%) / 6 | 3 (1%, 0,3–2,9%) / 3 | 3 (1%, 0,3–2,9%) / 3 | 1 (1%, 0,2–5,4%) / 1 | 1 (1%, 0,2–5,4%) / 1 | - |
|  | Unlikely | 2 (0,7%, 0,2–2,4%) / 3 | - | 2 (0,7%, 0,2–2,4%) / 3 | 1 (1%, 0,2–5,4%) / 1 | 1 (1%, 0,2–5,4%) / 1 | - |
|  | Unrelated | 4 (1,3%, 0,5–3,4%) / 9 |  | 4 (1,3%, 0,5–3,4%) / 9 | 3 (3%, 1,0–8,5%) / 8 | 1 (1%, 0,2–5,4%) / 1 | 2 (2%, 0,6–7,0%) / 7 |
|  | Unknown | 1 (0,3%, 0,1–1,9%) / 1 | 1 (0,3%, 0,1–1,9%) / 1 | - | 1 (1%, 0,2–5,4%) / 2 |  | 1 (1%, 0,2–5,4%) / 2 |
| Dizziness | Possible | 1 (0,3%, 0,1–1,9%) / 1 | - | 1 (0,3%, 0,1–1,9%) / 1 | - | - | - |
| **Musculoskeletal and connective tissue disorders** | Possible | 2 (0,7%, 0,2–2,4%) / 4 | 1 (0,3%, 0,1–1,9%) / 2 | 1 (0,3%, 0,1–1,9%) / 2 |  |  |  |
|  | Unlikely | 1 (0,3%, 0,1–1,9%) / 2 |  | 1 (0,3%, 0,1–1,9%) / 2 | 1 (1%, 0,2–5,4%) / 1 | 1 (1%, 0,2–5,4%) / 1 | - |
|  | Unrelated | 2 (0,7%, 0,2–2,4%) / 2 | 1 (0,3%, 0,1–1,9%) / 1 | 1 (0,3%, 0,1–1,9%) / 1 |  |  |  |
| Arthralgia | Possible | 2 (0,7%, 0,2–2,4%) / 2 | 1 (0,3%, 0,1–1,9%) / 1 | 1 (0,3%, 0,1–1,9%) / 1 | - | - | - |
|  | Unlikely | 1 (0,3%, 0,1–1,9%) / 1 |  | 1 (0,3%, 0,1–1,9%) / 1 |  |  |  |
| Lumbar pain | Unlikely | - | - | - | 1 (1%, 0,2–5,4%) / 1 | 1 (1%, 0,2–5,4%) / 1 | - |
| Myalgia | Possible | 2 (0,7%, 0,2–2,4%) / 2 | 1 (0,3%, 0,1–1,9%) / 1 | 1 (0,3%, 0,1–1,9%) / 1 | - | - | - |
|  | Unlikely | 1 (0,3%, 0,1–1,9%) / 1 |  | 1 (0,3%, 0,1–1,9%) / 1 |  |  |  |
|  | Unrelated | 2 (0,7%, 0,2–2,4%) / 2 | 1 (0,3%, 0,1–1,9%) / 1 | 1 (0,3%, 0,1–1,9%) / 1 |  |  |  |
| **Gastrointestinal tract disorders** | Possible | 2 (0,7%, 0,2–2,4%) / 2 |  | 2 (0,7%, 0,2–2,4%) / 2 |  |  |  |
|  | Unlikely | 2 (0,7%, 0,2–2,4%) / 2 | 1 (0,3%, 0,1–1,9%) / 1 | 1 (0,3%, 0,1–1,9%) / 1 |  |  |  |
|  | Unrelated |  |  |  | 1 (1%, 0,2–5,4%) / 1 | - | 1 (1%, 0,2–5,4%) / 1 |
| Diarrhea | Possible | 1 (0,3%, 0,1–1,9%) / 1 | - | 1 (0,3%, 0,1–1,9%) / 1 |  |  |  |
|  | Unrelated |  |  |  | 1 (1%, 0,2–5,4%) / 1 | - | 1 (1%, 0,2–5,4%) / 1 |
| Pyrosis | Unlikely | 1 (0,3%, 0,1–1,9%) / 1 | 1 (0,3%, 0,1–1,9%) / 1 | - | - | - | - |
| Nausea | Possible | 1 (0,3%, 0,1–1,9%) / 1 | - | 1 (0,3%, 0,1–1,9%) / 1 | - | - | - |
|  | Unlikely | 1 (0,3%, 0,1–1,9%) / 1 | - | 1 (0,3%, 0,1–1,9%) / 1 | - | - | - |
| **Hearing disorders** | Unlikely | 1 (0,3%, 0,1–1,9%) / 1 | 1 (0,3%, 0,1–1,9%) / 1 | - |  |  |  |
| Ear congestion | Unlikely | 1 (0,3%, 0,1–1,9%) / 1 | 1 (0,3%, 0,1–1,9%) / 1 | - | - | - | - |
| **Disorders of respiratory system and mediastinal organs** | Unlikely | 2 (0,7%, 0,2–2,4%) / 2 | 1 (0,3%, 0,1–1,9%) / 1 | 1 (0,3%, 0,1–1,9%) / 1 | 1 (1%, 0,2–5,4%) / 1 | 1 (1%, 0,2–5,4%) / 1 | - |
|  | Unrelated | 5 (1,7%, 0,7–3,9%) / 5 | 1 (0,3%, 0,1–1,9%) / 1 | 4 (1,3%, 0,5–3,4%) / 4 | 1 (1%, 0,2–5,4%) / 3 | - | 1 (1%, 0,2–5,4%) / 3 |
|  | Unknown |  |  |  | 1 (1%, 0,2–5,4%) / 1 | - | 1 (1%, 0,2–5,4%) / 1 |
| Throat pain | Unrelated | 2 (0,7%, 0,2–2,4%) / 2 | - | 2 (0,7%, 0,2–2,4%) / 2 | 1 (1%, 0,2–5,4%) / 2 | - | 1 (1%, 0,2–5,4%) / 2 |
| Pain in the oropharynx | Unrelated | 1 (0,3%, 0,1–1,9%) / 1 | - | 1 (0,3%, 0,1–1,9%) / 1 | - | - | - |
|  | Unknown |  |  |  | 1 (1%, 0,2–5,4%) / 1 | - | 1 (1%, 0,2–5,4%) / 1 |
| Cough | Unlikely | 1 (0,3%, 0,1–1,9%) / 1 | - | 1 (0,3%, 0,1–1,9%) / 1 |  |  |  |
|  | Unknown |  |  |  | 1 (1%, 0,2–5,4%) / 1 | - | 1 (1%, 0,2–5,4%) / 1 |
| Impaired sense of smell | Unrelated | 1 (0,3%, 0,1–1,9%) / 1 | 1 (0,3%, 0,1–1,9%) / 1 | - | - | - | - |
| Dyspnea | Unrelated | 1 (0,3%, 0,1–1,9%) / 1 | - | 1 (0,3%, 0,1–1,9%) / 1 | - | - | - |
| Sore throat | Unlikely | 1 (0,3%, 0,1–1,9%) / 1 | 1 (0,3%, 0,1–1,9%) / 1 | - | 1 (1%, 0,2–5,4%) / 1 | 1 (1%, 0,2–5,4%) / 1 | - |
| **Metabolic disorders** | Possible | 1 (0,3%, 0,1–1,9%) / 1 |  | 1 (0,3%, 0,1–1,9%) / 1 |  |  |  |
|  | Unlikely | 1 (0,3%, 0,1–1,9%) / 1 |  | 1 (0,3%, 0,1–1,9%) / 1 |  |  |  |
|  | Unrelated | 1 (0,3%, 0,1–1,9%) / 5 | - | 1 (0,3%, 0,1–1,9%) / 5 | 1 (1%, 0,2–5,4%) / 1 | - | 1 (1%, 0,2–5,4%) / 1 |
| Impaired appetite | Possible | 1 (0,3%, 0,1–1,9%) / 1 |  | 1 (0,3%, 0,1–1,9%) / 1 |  |  | - |
|  | Unlikely | 1 (0,3%, 0,1–1,9%) / 1 |  | 1 (0,3%, 0,1–1,9%) / 1 |  |  |  |
|  | Unrelated | 1 (0,3%, 0,1–1,9%) / 5 | - | 1 (0,3%, 0,1–1,9%) / 5 | 1 (1%, 0,2–5,4%) / 1 | - | 1 (1%, 0,2–5,4%) / 1 |
| **Other** | Unlikely | 1 (0,3%, 0,1–1,9%) / 1 | - | 1 (0,3%, 0,1–1,9%) / 1 |  |  |  |
| Death from acute circulatory disorder | Unlikely | 1 (0,3%, 0,1–1,9%) / 1 | - | 1 (0,3%, 0,1–1,9%) / 1 |  |  |  |
